# Cortical effects of dopamine replacement account for clinical response variability in Parkinson’s disease

**DOI:** 10.1101/2024.11.20.24317429

**Authors:** Alex I. Wiesman, Mikkel C. Vinding, Panagiota Tsitsi, Per Svenningsson, Josefine Waldthaler, Daniel Lundqvist

## Abstract

Individual variability in clinical response to dopaminergic replacement therapy (DRT) is a key barrier to efficacious treatment for patients with Parkinson’s disease (PD). A better understanding of the neurobiological sources of such inter-individual differences is necessary to inform future clinical interventions and motivate translational research. One potential source of this variability is an unintended secondary activation of extra-nigrostriatal dopamine systems by DRT, particularly in the neocortex. In this study, we use magnetoencephalography data collected from patients with PD before and after DRT to map their individual cortical neurophysiological responses to dopaminergic pharmacotherapy. By combining these DRT response maps with normative atlases of cortical dopamine receptor and transporter densities, we link the variable enhancement of rhythmic beta activity by DRT to dopamine-rich cortical regions. Importantly, patients who exhibited a larger dopaminergic beta cortical enhancement showed a smaller clinical improvement from DRT, indicating a potential source of individual variability in medication response for patients with PD. We conclude that these findings inform our understanding of the dopaminergic basis of neurophysiological variability often seen in patients with PD, and indicate that our methodological approach may be useful for data-driven contextualization of medication effects on cortical neurophysiology in future research and clinical applications.

## Introduction

To reduce movement symptoms associated with Parkinson’s disease (PD), the vast majority of patients are prescribed dopamine replacement therapy (DRT) such as levodopa. Even across pathologically-confirmed cases of PD, the magnitude and nature of motor symptom improvement from dopaminergic medications can vary substantially, with as many as 25% of such patients being considered non-responsive to levodopa therapy pre-mortem^1^. Such variability in patient responses to these medications represents a key challenge, not only for effective symptom management across patients, but also for early and accurate detection, as DRT responsiveness is considered a key feature in the differential diagnosis of PD^2^. A more nuanced understanding of the variable neurophysiological effects of dopaminergic pharmacotherapies may help shed light on the source of clinical response variability to these medications.

Dopaminergic medications aim to rescue motor function in PD via restoration of dopaminergic signaling in the nigrostriatal circuit^3^. This restored signaling manifests at the level of the cortex as normalized rhythmic beta-frequency (15 – 30 Hz) neurophysiology in primary motor regions, leading to improved motor function^4,5^. However, the effects of dopamine medications on neurophysiology are widespread, as they do not specifically activate dopamine receptors in the basal ganglia. Levodopa and related medications have been shown to exert effects on neurophysiology across the cortex, both within and outside of the beta frequency range^5–8^. Such effects on cortical neurophysiology may partially explain inter-individual variability in motor^1^ and non-motor^9^ DRT response across patients with PD. Yet, difficulties in parsing the effects of DRT on cortical neurophysiology via the unintended activation of cortical dopamine receptors versus via the intended modification of nigrostriatal signaling has hindered the study of these unintended effects.

To address this issue, we pair pharmaco-magnetoencephalography (MEG) with an atlas of cortical dopamine system densities to link DRT effects on cortical beta activity to cortical dopamine circuits in a group of patients with PD. Using a partial least squares framework, we find that the administration of DRT enhances beta-frequency rhythmic activity in regions of the cortex with high densities of dopamine receptors and transporters in some patients, but not in others. Importantly, the magnitude of this beta-dopamine enhancement is associated with reduced clinical responses to DRT, indicating that secondary activation of cortical dopamine systems by dopamine pharmacotherapies may account for response variability in patients with PD. We anticipate that these findings will be useful in improving personalized pharmacological regimens for patients with PD. Our analytical approach may also prove valuable for future research into neurophysiological sources of inter-individual variability in other neurological and psychiatric disorders.

## Methods

### Participants

MEG data from 17 patients with mild-to-moderate idiopathic PD (Hoehn and Yahr stages 1 – 3; age 41 – 85; 5 female) and 20 healthy older adults (age 54 – 76; 8 female) were re-analyzed from a previous study of dopamine therapy effects on neurophysiological activity in PD^10,11^. These groups were matched in terms of age and self-reported sex (Table 1). Participants provided written informed consent prior to enrollment in the study, and all protocols complied with the Declaration of Helsinki. The regional research ethics committee in Stockholm reviewed and approved this study (DNR 2016/19-31/1 Regionala Etikprövningsnämnden Stockholm, latest amendment: DNR 2024-00658-02, The Swedish Ethical Review Authority). Non-demented patients with PD were recruited who were prescribed a stable and effective DRT regimen, who had been diagnosed with PD based on the Movement Disorder Society clinical diagnostic criteria for PD^12^, and who had no other major neurological or psychiatric diagnoses. Levodopa equivalent daily dosage (LEDD) was calculated for each patient following a standard approach^13^. Details on the patient group’s clinical profile are provided in Table 2.

**Table 1.**
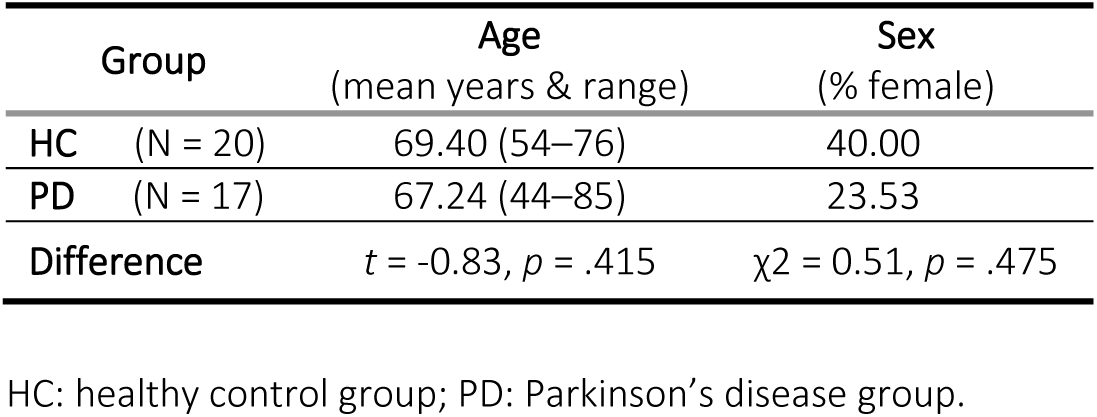
Demographic group comparisons.

| Group |  | Age<br>(mean years & range) | Sex<br>(% female) |
| --- | --- | --- | --- |
| HC | (N = 20) | 69.40 (54–76) | 40.00 |
| PD | (N = 17) | 67.24 (44–85) | 23.53 |
| Difference | | $t = -0.83, p = .415$ | $\chi^2 = 0.51, p = .475$ |

HC: healthy control group; PD: Parkinson’s disease group.

**Table 2.** Patient group clinical profile.

|  | UPDRS-III<br>OFF | UPDRS-III<br>ON | Hoehn &<br>Yahr** | MoCA | Disease Duration<br>(years)* | LEDD† |
| --- | --- | --- | --- | --- | --- | --- |
| **Range** | 10–61 | 5–39 | 1–3 | 19–29 | 1–11 | 300–1150 |
| **Mean (SD)** | 33.06 (11.77) | 17.41 (9.70) | 2.27 (0.59) | 25.24 (2.95) | 5.31 (3.94) | 634.69 (250.70) |

UPDRS-III: Unified Parkinson's Disease Rating Scale part III; OFF/ON: before/after administration of patient's normal levodopa regimen; LEDD: levodopa equivalent daily dosage; MoCA: Montreal Cognitive Assessment. \*Data missing for one patient. \*\*Data missing for two patients.

### MEG & MRI data acquisition

For each MEG recording session, data were acquired at a sampling rate of 1kHz with online 0.1 Hz high-pass and 330 Hz low-pass filters using an Elekta Neuromag TRIUX 306-channel system inside of a two-layer magnetically shielded room (Vacuumschmelze GmbH, model Ak3b). During recording, participants sat with their eyes open and fixated on a centrally presented crosshair for 8 minutes. Continuous head position indicator coils were used to track the movements and position of each participant throughout the session. The position of these coils, along with approximately 80 points spaced across the scalp, were digitized using a Polhemus FASTRAK (Polhemus Inc.) system prior to MEG recordings for offline coregistration of the MEG data to the participant-specific structural MRIs for source imaging. Electro-oculogram (EOG) and electrocardiogram (ECG) data were recorded simultaneously for correction of ocular and heartbeat artifacts, respectively.

All participants underwent two such MEG recording sessions. For the patients with PD, the first of these sessions occurred after at least 12 hours of withdrawal from their normal DRT regimen – that is, in the practically-defined OFF state. After this, patients took their normal medication and waited one hour before another identical MEG recording session in the ON state. Healthy older adults also underwent two MEG recording sessions one hour apart, to control for any potential effects of time and/or familiarity with the scanning environment. Clinical motor function was measured by a trained neurologist using the Movement Disorder Society’s Unified Parkinson’s Disease Rating Scale – part III (UPDRS-III) immediately after each MEG session for patients with PD.

Structural T1-weighted MRI data were acquired for each participant on either a GE Discovery 3T or a Siemens Prisma 3T system. Note that scanner differences are unlikely to affect the accuracy of MEG source imaging, which was the only use case for the MRI data in the present study. One participant did not complete an MRI.

### MEG data processing

Temporal signal space separation (tSSS; window = 10 s, correlation limit = 0.95)^14^ and movement compensation (median head position) were applied to the MEG data, which were then downsampled to 200 Hz to facilitate processing and low-pass filtered at 100 Hz. Independent component analysis (ICA) was used to remove ocular and cardiac artifacts from the data with the *fastica* algorithm^15^, based on their component correlations with the EOG/ECG signals. Source reconstruction was performed using dynamic statistical parametric mapping (dSPM)^16^ with a noise covariance matrix estimated using 2 minutes of empty-room data collected prior to each session. The inner skull boundary of each participant’s MRI was used to generate a single compartment volume conductor forward model. Individual anatomical surfaces with 5124 evenly-spaced locations were generated following segmentation of the MRI data using *recon-all* in freesurfer^17^ (version 5.3). For one participant who did not provide MRI data, a pseudo-individualized surface was generated by warping an MRI template^18^ to their digitized head surface points.

The cortical surface was parcellated using the Desikan-Killiany atlas^19^ and continuous data extracted from each of the 68 regions of interest by taking the first right-singular vector of a singular value decomposition of it’s constituent source time series (sign normalized relative to source orientations). These region-wise time series were then transformed into power spectral densities (PSD) using Welch’s method (time windows = 2.56 seconds; 50% overlap). Pre-processing, source imaging, and frequency transformation of the data were performed in MNE-Python^20^ (Python version 3.6).

To disentangle neurophysiological effects due to rhythmic versus arrhythmic signaling, we parameterized the PSDs with specparam (Python implementation; frequency range = 2–40 Hz; Gaussian peak model; peak width limits = 0.5 –12 Hz; maximum n peaks = 3; minimum peak height = 0.1; fixed aperiodic). The arrhythmic component of the power spectra was represented by the aperiodic slope and the rhythmic spectra were represented by subtracting the arrhythmic power spectra from the original PSDs and then averaging over canonical frequency bands (delta: 2–4 Hz; theta: 5–7 Hz; alpha: 8–12 Hz; beta: 15–30 Hz). This resulted in maps of region-wise neurophysiological activity per session and participant for five features: one for the arrhythmic slope and each of the four rhythmic frequency bands. Based on previous research, our primary analyses focused on alpha^8,21–23^ and beta frequency rhythmic activity^5,8,11,24,25^, as well as arrhythmic neurophysiology^10,11,26–32^, but supplementary analyses also investigated the importance of delta and theta rhythmic signaling.

### Modeling DRT-responses and their spatial alignment to cortical dopamine systems

To generate maps of DRT-response per each of the neurophysiological features (Fig. 1a), we first subtracted the first session maps of each feature from the second session maps. In patients with PD, this resulted in a difference between their ON-and OFF-DRT sessions, while for healthy adult participants, it resulted in a difference representing any effects of the session order (e.g., time of day, familiarity with the MEG environment). The ON-minus-OFF maps for each patient were then standardized (i.e., z-scored) relative to maps of the mean and standard deviation of comparable differences in the healthy adult group. This generated maps for each patient with PD per feature representing the effects of DRT, corrected for session effects.

**Figure 1.**
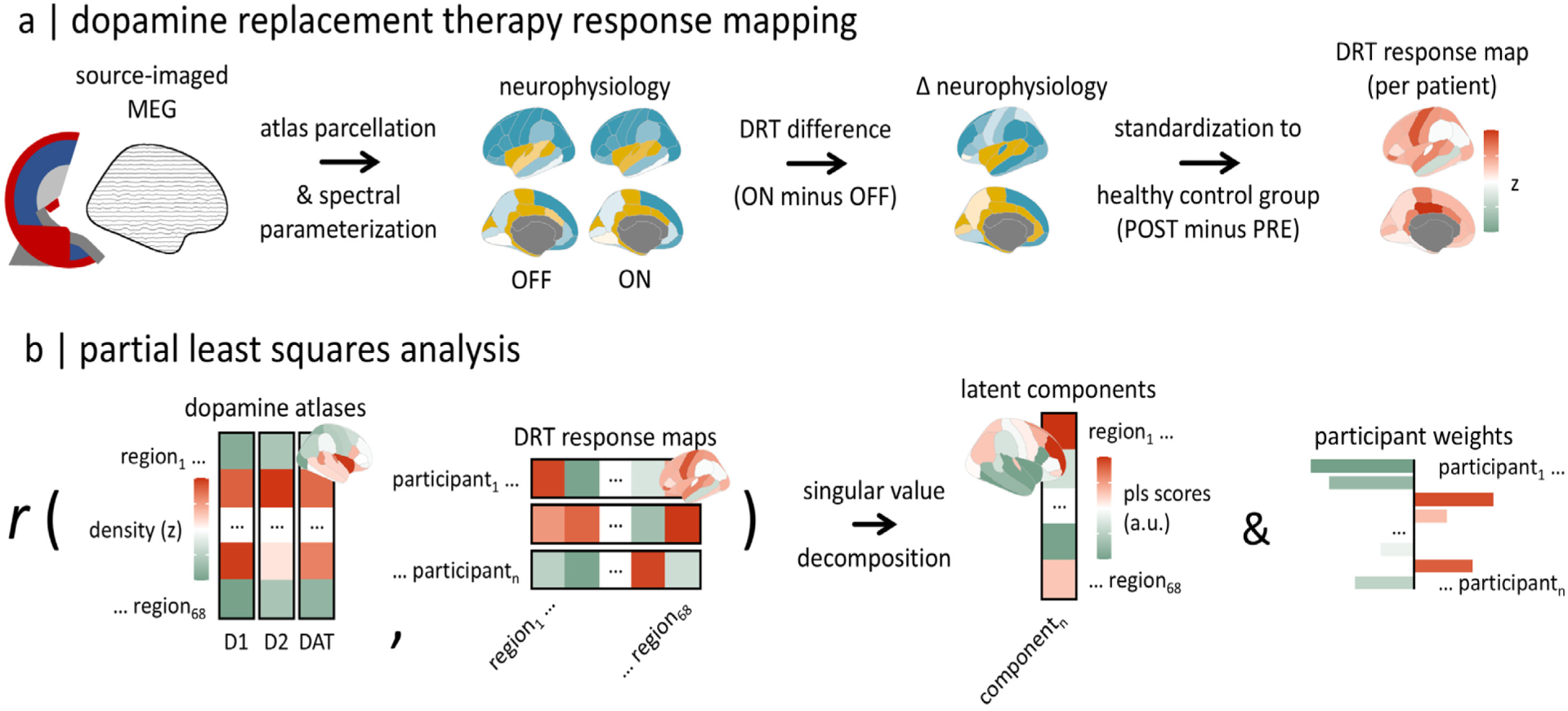
Mapping neurophysiological responses to dopamine replacement therapy and neurochemical alignment analysis. (a) To generate maps of neurophysiological response to dopamine replacement therapy (DRT), we parcellated preprocessed and source-imaged magnetoencephalography (MEG) data to the Desikan-Killiany atlas and used *specparam* to separate rhythmic from arrhythmic patterns of signaling. This was done for both the PRE and POST (healthy controls; HC) or OFF and ON medication (patients with Parkinson’s disease; PD) timepoints, and these maps were then subtracted to derive Δ neurophysiology maps for each participant (ON-OFF for PD and POST-PRE for HC). The ON-OFF map for each patient was then standardized (i.e., z-scored) to mean and standard deviation maps of the POST-PRE data from the HC group, to generate DRT response maps representing the effect of dopamine medications, controlling for any effects of timepoint. This procedure was repeated for each neurophysiological feature of interest, i.e., the delta, theta, alpha, and beta rhythmic signaling maps, as well as the arrhythmic signaling maps generated with the aperiodic slope. (b) These DRT response maps were then used as predictors in a partial least squares analysis to determine their spatial alignment to three normative atlases of dopamine system densities from *neuromaps*. Latent components (i.e., pls scores) from this analysis represent the spatial pattern of dopamine system densities that align most strongly with DRT neurophysiological responses, and the participant weights represent the strength by which each patient’s DRT response matches that pattern. Stringent autocorrelation-preserving null models and multiple comparisons corrections were used to ensure conservative inferences.

Neurochemical atlases were derived, as in several previous studies, from *neuromaps*^26,33–36^. Dopamine atlases included averaged positron emission tomography (PET) maps of D1 receptor densities measured from 13 adults (mean age = 33.00 years) using [11C]SCH23390 PET, D2 receptor densities measured from 92 adults (mean age = 38.89 years) using [11C]FLB-457 PET, and dopamine transporter (DAT) densities measured from 174 adults (mean age = 61.00 years) using [123I]-FP-CIT PET. Due to known associations between alpha rhythmic activity and norepinephrine signaling in PD^21^, we also extracted an atlas of norepinephrine transporter (NET) densities, measured from 77 adults using [11C]MRB PET.

These DRT response maps and dopamine atlases were used in a partial least squares regression (PLS)^37,38^ to examine the spatial alignment of DRT effects on neurophysiology to dopamine systems across patients (Fig. 1b). Specifically, our PLS performed a dual decomposition of two matrices – the 68 region-by-17 patient matrix of DRT response maps and the 68 region-by-3 PET tracer (i.e., D1, D2, and DAT) matrix of dopamine atlases – such that the derived components explained the maximum amount of spatial covariance between these matrices. In the case of our study, this served to identify patterns of spatial alignment between neurophysiological responses to DRT and cortical dopamine receptor/transporter densities that are variable across patients and may otherwise be averaged out in traditional statistical analysis. Importantly, PLS is also robust to high collinearity between variables, such as is present between the three dopamine atlases used in our analysis. The statistical significance of the resulting DRT response-dopamine alignments (i.e., the first PLS component) was determined by testing the variance explained by each against a null distribution of explained variances derived from 5,000 similar PLS models that we computed with autocorrelation-preserving spatial permutations^39^ of the dopamine atlases. Similar models were computed with alpha and beta-frequency DRT response maps using the NET atlas to examine potential alignments to norepinephrine systems^21^. Multiple comparisons across PLS models for each neurotransmitter system were controlled for the false discovery rate by applying the Benjamini-Hochberg procedure to the resulting non-parametric p-values, with a final significance threshold of *p*_FDR_ < .05. PLS analysis was performed using the *plsr*^40^ package in *R*.

### Modeling clinical associations of DRT responses in dopamine-rich regions

From the neurochemical PLS models, loading weights were extracted to represent the strength with which each patient exhibited the significant pattern of neurophysiological response in dopamine-rich cortical regions. Higher values for these weights indicate that a patient’s DRT response was stronger in cortical regions with high densities of dopamine receptors/transporters, while lower values indicate the opposite. These weights were regressed on the ON-minus-OFF ΔUPDRS-III scores, to test whether the cortical neurophysiological activation of dopamine systems by DRT was related to clinical responses to DRT. Age, sex, and symptom laterality were considered as potential nuisance covariates, with step-wise model comparison based on Akaike Information Criterion (PLS weights as dependent variable; threshold of ΔAIC > 2; both directions) used to select nuisance regressors for the final model. Similar regressions were computed using the ON and OFF UPDRS-III scores separately, to ensure that any relationships with the ΔUPDRS-III scores were not due to confounds (e.g., patients with higher UPDRS-III OFF scores having more room to improve). A secondary model was also computed with LEDD as a covariate, to determine whether any significant relationships could be explained by the relative dopaminergic impact of DRT.

## Results

### Clinical and neurophysiological responses to dopamine replacement therapies

We examined the clinical and neurophysiological responses to DRT in patients with idiopathic PD (Fig. 1a). As expected, patients with PD exhibited a strong clinical response to DRT (mean ON-OFF ΔUPDRS-III = - 15.65; mean % decrease in UPDRS-III = 49.35) with notable variability across individuals (range = 25.58% – 72.73%, standard deviation = 14.97%). These patients also showed cortical neurophysiological responses to DRT in rhythmic beta (Fig. 2a) and alpha (Fig. S1) activity, with less robust responses in arrhythmic (i.e., aperiodic) signaling (Fig. S1). Intriguingly, and unlike in the cases of arrhythmic and alpha rhythmic activity, many of the cortical regions that exhibited the most variable patterns of rhythmic beta responses to DRT were also the areas with the highest densities of dopamine receptors and transporters (Fig. 2b).

**Figure 2.**
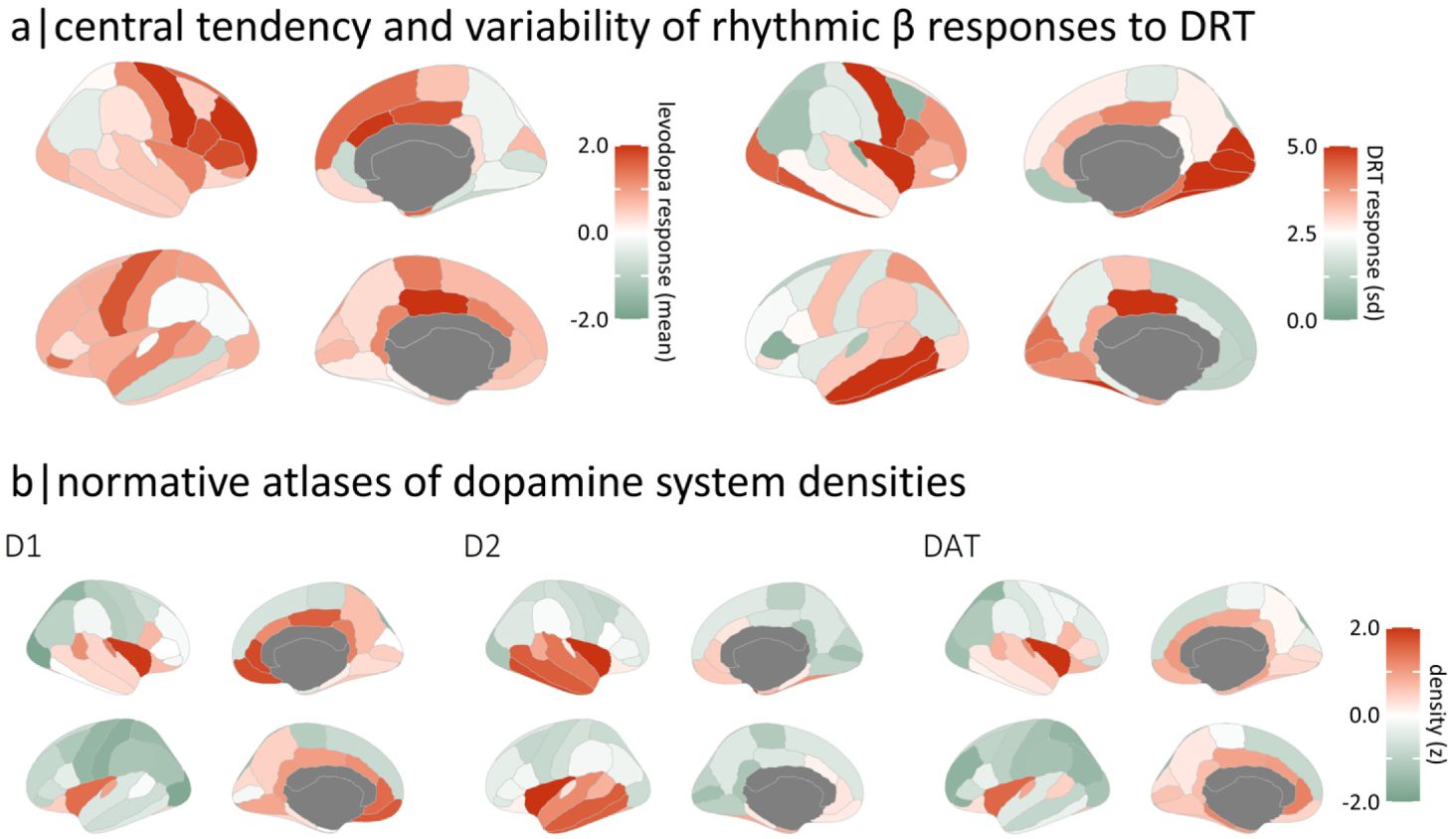
Rhythmic beta-frequency responses to dopamine replacement therapy. (a) Brain maps on the left and right indicate the mean and standard deviation, respectively, of region-wise rhythmic beta-frequency neurophysiological responses to dopamine replacement therapy (DRT). (b) From left to right, brain maps indicate the normative spatial patterns of D1 and D2 receptor and dopamine transporter densities.

### Variable beta rhythmic activation of dopaminergic cortical areas by dopamine replacement therapies

We next tested whether these variable responses to DRT in beta rhythmic activity occurred in cortical regions with high densities of dopamine receptors and transporters (Fig. 1b). Inter-individual variability in rhythmic beta-frequency neurophysiological responses to DRT was aligned with regions of high dopamine densities (Fig. 3a; first component; D1: 18.15% spatial variance explained; D2: 29.36%; DAT: 24.95%; *p*_FDR_ = .009). This alignment was such that, in some patients with PD, DRT increased rhythmic beta-frequency signaling in inferior frontal and temporal cortices rich in dopamine receptors and transporters, while in others, no such increase occurred (Fig. 3b, left). No significant alignments were observed between dopamine systems and delta, theta, or alpha rhythmic responses to DRT (all *p*_FDR_ > .400), between dopamine systems and arrhythmic responses to DRT (*p*_FDR_ = .200), nor between norepinephrine densities and alpha or beta rhythmic responses (all *p*_FDR_ > .150).

**Figure 3.**
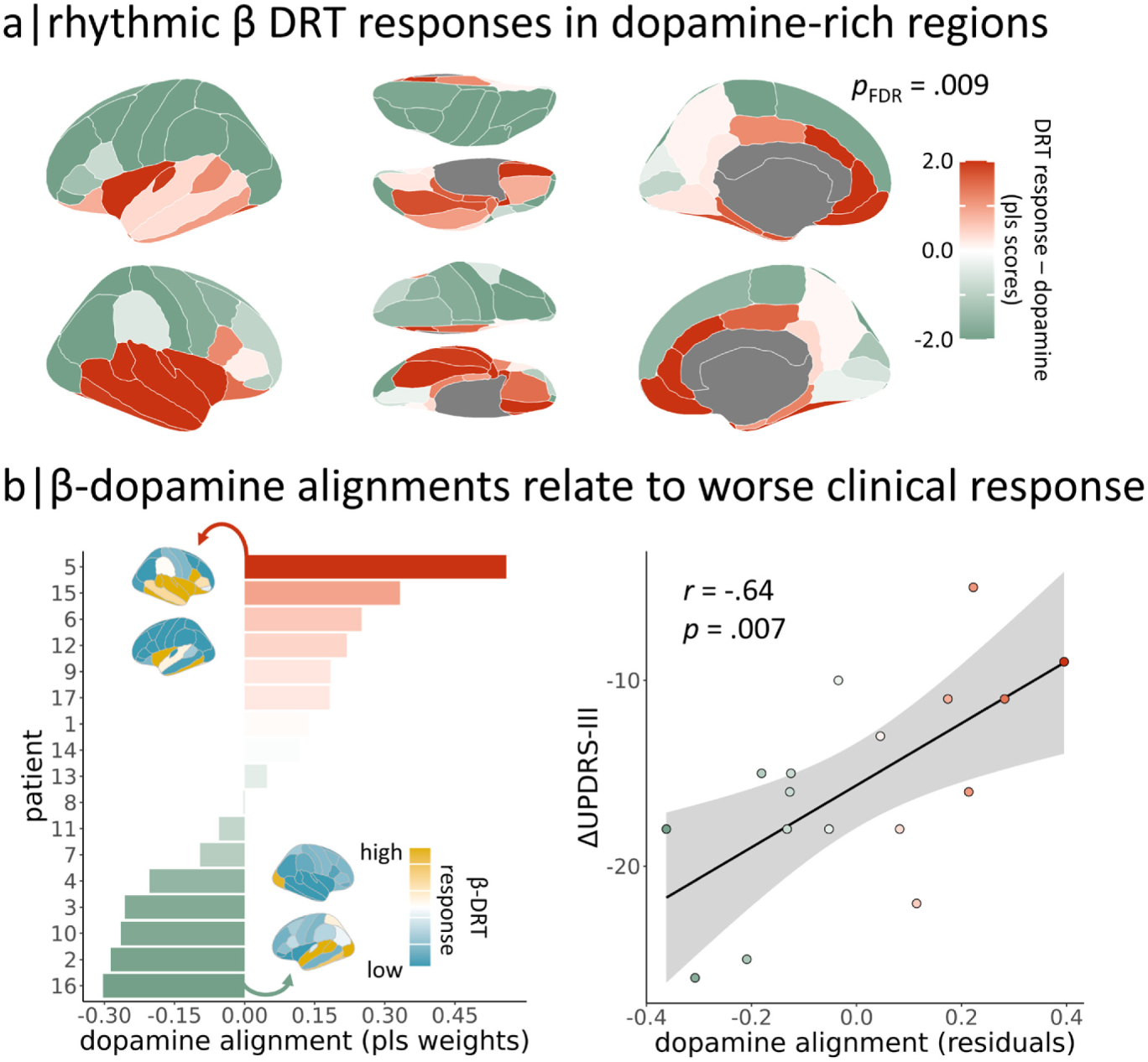
Rhythmic beta-frequency responses to dopamine replacement therapy align with cortical dopamine systems and relate to clinical response. (a) Brain maps indicate the spatial pattern of dopamine system densities that most strongly aligned with variable rhythmic beta-frequency responses to dopamine replacement therapy (DRT). This alignment was significant (*p*_FDR_ = .009) following stringent autocorrelation-preserving spatial permutation and multiple comparison corrections. (b) Those patients with Parkinson’s disease (left, y-axis) who exhibited stronger rhythmic beta responses to DRT in dopamine-rich cortices (left and right, x-axes) showed less improvement in motor function from DRT (right, y-axis). Brain maps on the left plot are rhythmic beta responses to DRT from representative patients with the highest (top) and lowest (bottom) such alignment.

### Clinical response to dopamine replacement therapies is related to the activation of dopaminergic cortices

Stronger beta-rhythmic responses to DRT in dopamine-rich cortical regions were associated with worse clinical responses to DRT across patients (Fig. 3b, right; *r* = -.64, *p* = .007), beyond the effects of symptom laterality (for details regarding covariate selection, see *Methods: Modeling clinical associations*). Importantly, this relationship was specific to clinical responses to DRT (i.e., ON-OFF changes in the UPDRS-III), and did not replicate when considering only the ON (*p* = .826) or OFF (*p* = .337) motor symptom scores. It also remained significant when LEDD was included in the model (*r* = -.57, *p* = .032). Independent variance in this beta-dopamine effect was also intuitively linked to symptom laterality (*r* = -.62, *p* = .011) beyond its relationship to clinical response, such that patients exhibited stronger beta-rhythmic responses to DRT in primary motor cortices contralateral to their more-affected side.

## Discussion

Quantitative markers of medication response variability promise to both improve therapeutic interventions and enhance basic understanding of pathophysiology in PD. Neurophysiological measurements with non-invasive imaging methods are dynamic and repeatable, making them a strong candidate. In this study, we find that the individual whole-cortex spatial patterns of neurophysiological responses to DRT measured by MEG can be used to delineate unintended activation of cortical dopaminergic systems in patients with PD. These activations are then associated with worse medication responses across individuals, implicating cortical dopamine signaling alterations in clinical DRT response variability in PD.

We find *beta-frequency rhythmic activity* to be specifically relevant. First, the inter-individual variation in enhancement of beta rhythmic activity after DRT is aligned with dopamine-rich regions of the cortex. Second, the magnitude of these variable dopamine-beta responses is associated with symptom severity, so that a large beta enhancement is associated with less motor function improvement. Our results agree with decades of previous research connecting beta oscillations to the pathophysiology of PD^25^. Increased beta coherence in the nigrostriatal circuit is implicated in worsened motor symptoms in PD^25,41–44^, and is normalized by therapeutics^45–50^. This beta-frequency increase appears to be propagated to local somato-motor cortical activity in the early stages of the disease^51^, but then shifts to a marked reduction in the later stage^5,11,21,26,52,53^. Increase of motor-cortical beta activity with dopamine medications in these later stages is then associated with motor symptom improvement^6,48^.

As such, our findings suggest an important clinical distinction between primary beta-rhythmic activation of cortical dopamine systems versus beta-rhythmic changes secondary to normalization of nigrostriatal signaling. Patients in whom DRT elicits beta rhythmic increases in somato-motor regions, where nigrostriatal pathways exert influence on the cortex and control movement, are more clinically responsive. This effect is particularly pronounced in the hemifield corresponding to the patients’ more affected side. In contrast, those who exhibit beta rhythmic increases in non-somato-motor cortical regions that are rich in dopamine systems have less pronounced motor symptom improvements from DRT.

No significant alignments were found between alpha rhythmic responses to DRT and either dopamine or norepinephrine systems. Though alpha-frequency increases in response to DRT have been reported^8^, we find no evidence here that these responses are focused in brain regions dense in dopamine systems, suggesting that alpha rhythmic changes resultant of DRT are not due to primary activation of cortical dopamine receptors. This also supports recent work showing that alpha-frequency alterations seen in patients with PD are instead linked to degeneration of the locus coeruleus and associated changes in noradrenergic cortical signaling^21^, which would not be expected to be impacted by DRT.

Our spectral parameterization approach adds nuance to these findings. Although arrhythmic neurophysiology is altered in PD^10,26–32^, there is mixed evidence as to whether it is affected by DRT^10,29,54^. We did not observe strong modulation of the 1/f arrhythmic exponent by DRT, nor high variability in any such modulation across patients. In contrast, 1/f-corrected rhythmic activity in the alpha and beta frequencies exhibited substantial DRT response variability, both across patients and brain regions, and was aligned with dopamine systems. This discrepancy indicates the value of separating rhythmic from arrhythmic neurophysiology when studying Parkinson’s disease and other neurological disorders.

A key limitation to our work is the lack of individual maps of cortical dopamine receptor and transporter densities. Although the current method makes a significant contribution to our understanding of the relationship between individual variation in response to DRT and dopamine receptor and transporter densities, individual maps would help to directly examine the relationship between the individual variation in response to DRT therapy and the inter-individual variations in receptor and transporter densities, so this is a clear next direction for our work. Another limitation of this work is the lack of detailed cognitive testing data in both the ON and OFF medication states. Cortical dopamine effects are thought to be relevant for cognitive difficulties in patients with PD^55–57^, and this is therefore another future direction for our work. The relatively small sample of participants also limits our ability to thoroughly investigate inter-individual variability in this work. However, it should be noted that our within-subjects contrast of ON versus OFF medication states reduces concerns over noise and increases statistical power. Regardless, similar work in a larger and more heterogeneous group of participants is essential to ensure generalizability and examine the moderations of these relationships by demographic and other clinical factors.

In sum, we find a pattern of cortical rhythmic beta activity elicited by DRT in patients with PD, distinct from that of nigrostriatal effects, which maps on to cortical dopamine systems and relates to individual variability in clinical response. We expect this pattern to be valuable for future research and clinical interventions where tracking unintended cortical DRT effects on dopamine systems is needed. We also foresee that our analytical approach of using a partial least squares framework to map variable spatial maps of brain activity onto neurochemical systems may be useful for future neurological and psychiatric research.

## Data Availability

The datasets collected for the current study contains patient information that cannot be made public. The dataset is available from the authors for review purpose or on reasonable request.

## Acknowledgements

This work was supported to AIW by a Banting Postdoctoral Fellowship (BPF-186555) and the Canada Research Chair (CRC-2023-00300) in Neurophysiology of Aging and Neurodegeneration from the Canadian Institutes of Health Research (CIHR) and grant F32-NS119375 from the United States National Institutes of Health (NIH). The original study, including the work of MCV, JW, and DL was supported by the Swedish Foundation for Strategic Research (SBE 13-0115). The NatMEG facility, at which these data were collected, is supported by the Knut and Alice Wallenberg Foundation (KAW2011.0207). The funders had no role in study design, data collection and analysis, decision to publish, or preparation of the manuscript.

## Supplementary Material

**Figure S1.**
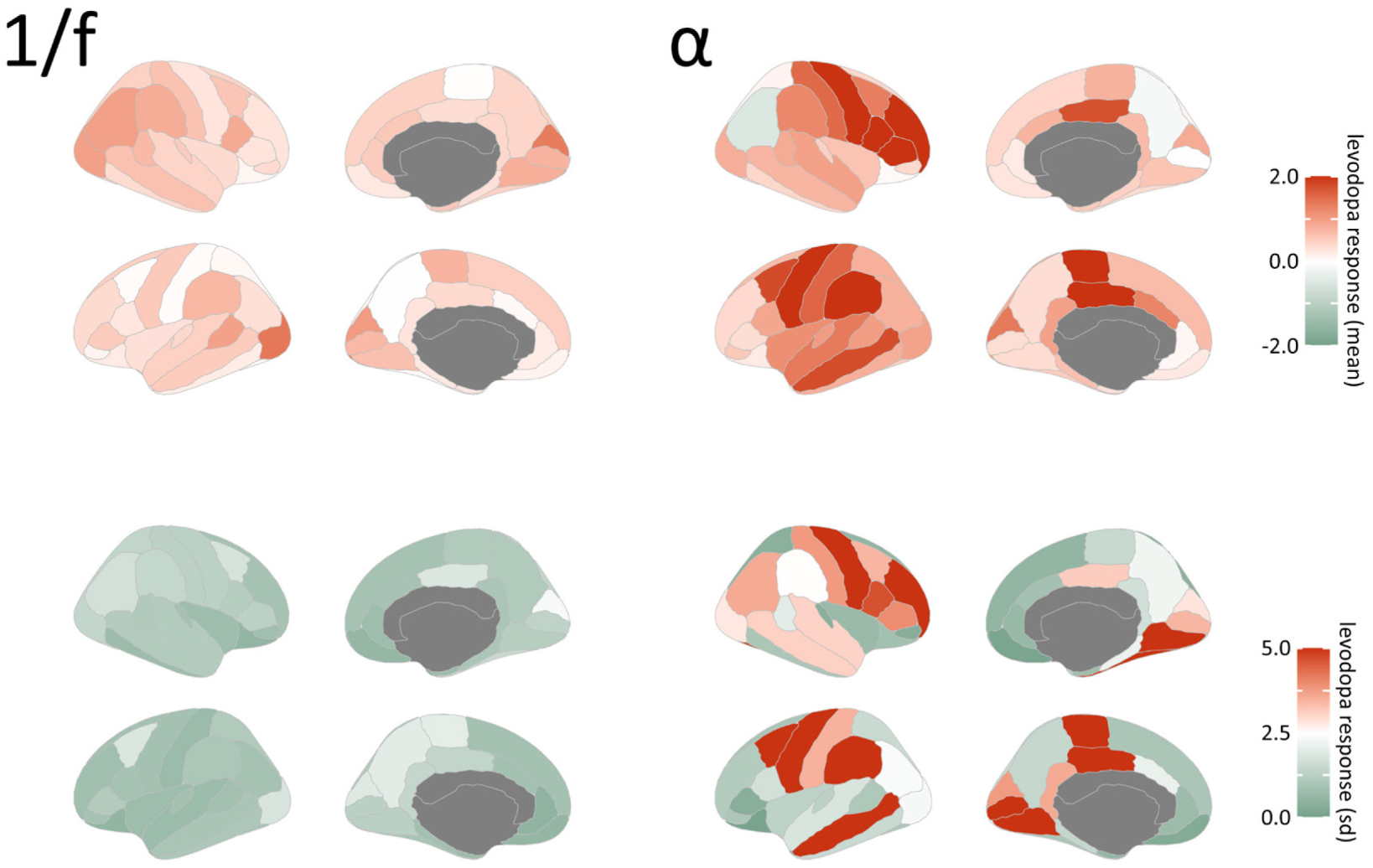
Arrhythmic and rhythmic alpha-frequency responses to dopamine replacement therapy. Similar to Figure 2, but for arrhythmic (i.e., 1/f or aperiodic exponent; left) and alpha-frequency rhythmic (right) neurophysiological activity.

